# Obesity shapes selection for driver mutations in cancer

**DOI:** 10.1101/2024.01.10.24301114

**Authors:** Cerise Tang, Venise Jan Castillon, Michele Waters, Chris Fong, Tricia Park, Sonia Boscenco, Susie Kim, Nikolaus Schultz, Irina Ostrovnaya, Alexander Gusev, Justin Jee, Ed Reznik

## Abstract

Obesity is a leading risk factor for cancer, but whether obesity is linked to specific genomic subtypes of cancer is unknown. Here, we examined the relationship between obesity and tumor genotype in two large clinicogenomic corpora. Obesity was associated with specific driver mutations in lung adenocarcinoma, endometrial carcinoma, and cancers of unknown primary, independent of clinical covariates and genetic ancestry. Obesity is therefore a putative driver of etiologic heterogeneity across cancers.

## Main

The physiologic and environmental forces leading to cancers with specific genotypes are largely unknown. Even as oncogenic mutations emerge in healthy tissue, a confluence of physiologic, immunologic, and epigenetic changes ultimately elicit tumorigenesis^1^. Obesity is a risk factor for the development of numerous cancers,^2,3^ and growing evidence suggests that medical interventions to reduce obesity can reduce cancer risk^4^. Obesity itself is associated with changes in systemic immune surveillance, metabolism, and inflammation^5–7^. In composite, these changes have the potential to shape the selective pressure for specific driver mutations in cancer, connecting the evolution of tumor-intrinsic genotypes to aspects of systemic health.

Testing the hypothesis that body mass index (BMI, a quantitative surrogate of obesity status) associates with specific tumor genotypes at population-scale necessitates the joint collection of clinical and genomic data that has been largely absent in large-scale clinical sequencing datasets to date. To overcome this gap, we systematically extracted information on body mass index (BMI) and other demographic factors in 34,274 patients profiled as part of their routine clinical care by the MSK-IMPACT clinical sequencing platform. We assessed the statistical association between BMI and tumor genotype by modeling the incidence of mutations in 341 cancer-associated genes as a function of BMI, focusing on gene-cancer type pairs with sufficient patient number for adequate statistical power (see **Methods**). Considering only oncogenic mutations as annotated by an FDA-recognized database^8^, we identified six genes across three separate cancer types demonstrating statistically significant enrichment with BMI in specific cancer types (q-value < 0.05) (**Figure 1a, Supplemental Table 1**). In lung adenocarcinoma, three genes (*KRAS* q-value = 2.6×10^−5^ estimate = 0.03, *SETD2* q-value = 1.31×10^−2^ estimate = 0.06, and *PPP2R1A* q-value = 1.31×10^−2^ estimate = 0.16) were mutated at higher frequency in obese patients, and one (*EGFR* q-value 3.0×10^−10^ estimate = −0.05) was mutated at lower frequency in obese patients, although the number of patients with *SETD2* and *PPP2R1A* mutations was small (**Table 1)**. In addition, we found *BAP1* in cancers of unknown primary to be positively associated with BMI (q-value *=* 3.5×10^−2^ estimate = 0.15) and *POLE* in uterine endometrioid carcinoma to be negatively associated with BMI (q-value = 1.6×10^−2^ estimate = −0.08). In contrast, we found no statistically significant associations between the presence of silent mutations (under putatively neutral selection) and BMI at the gene-cancer type level (q-value < 0.05) (**Supplemental Figure 1**).

**Table 1:** Number of patients per cancer-genotype and BMI category for statistically significant associations.

|  | *EGFR*<br>LUAD | *KRAS*<br>LUAD | *SETD*<br>2<br>LUAD | *PPP2R1A*<br>LUAD | All<br>LUAD | *BAP1*<br>CUP | All CUP | *POLE*<br>ENDO | All<br>ENDO | All<br>Patients |
| --- | --- | --- | --- | --- | --- | --- | --- | --- | --- | --- |
| Underweight | 39 | 26 | 3 | 0 | 113 | 0 | 14 | 2 | 8 | 1259 |
| Healthy | 513 | 456 | 41 | 1 | 1568 | 2 | 168 | 17 | 162 | 11957 |
| Overweight | 378 | 519 | 34 | 5 | 1499 | 4 | 139 | 23 | 205 | 11564 |
| Obese | 193 | 372 | 54 | 5 | 970 | 6 | 102 | 20 | 433 | 9494 |

LUAD = lung adenocarcinoma
CUP = cancer of unknown primary
ENDO = uterine endometrioid carcinoma

**Fig. 1:**
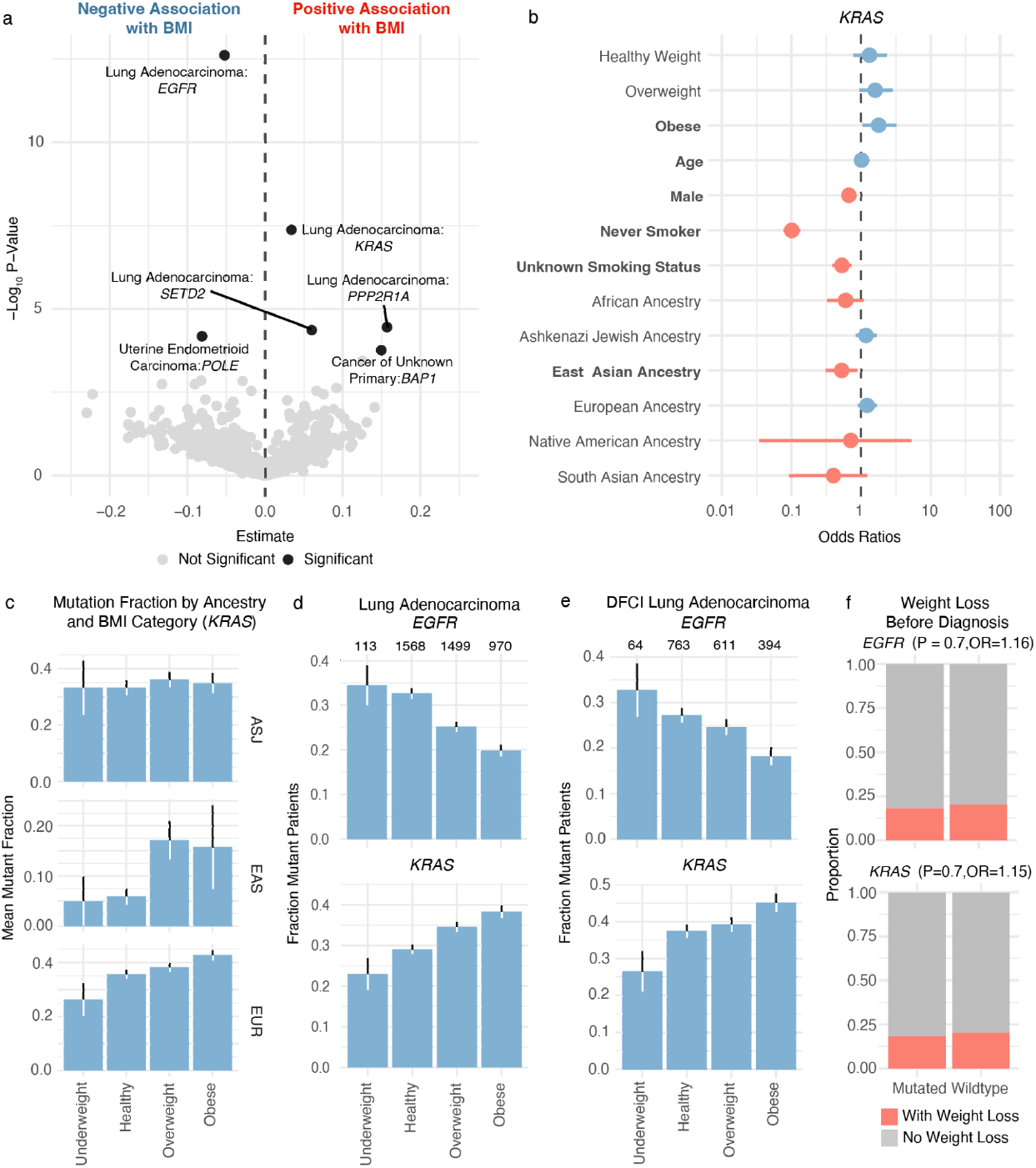
Oncogenic mutations are associated with body mass index. a) Statistical association between continuous BMI and genotype across gene-cancer-type pairs. −log_10_ p-values and estimates sizes from univariate logistic regression on the y and x-axis, respectively. Statistically significant pairs in black. b) Multivariate regression demonstrates that BMI categories are associated with *KRAS* mutations independent of other clinical factors. Result for multivariate regression with BMI as a continuous variable is in **Supplemental Table 3**. c) *KRAS* mutation frequency in lung adenocarcinoma patients categorized by BMI and genetic ancestry. ASJ = Ashkenazi Jewish. EAS = East Asian. EUR = European. d) *EGFR* (top) and *KRAS* (bottom) mutation frequency in BMI categories in MSKCC cohort. e) *EGFR* (top) and *KRAS* (bottom) mutation frequency in BMI categories in DFCI cohort. f) *EGFR* (left) and *KRAS* (right) are not associated with pre-diagnosis weight loss in lung adenocarcinoma.

Driver mutations are known to accumulate at different rates in patients of distinct genetic ancestries, raising the possibility that an association between tumor genotypes and obesity status could emerge indirectly^9^. To control for the contributions of genetic ancestry, we obtained ancestry information for each patient from a previously reported computational analysis of ancestry from germline sequencing data^10^. After correcting for age, sex, and genetic ancestry, all six significant univariate associations between BMI and genotype remained statistically significant in the same direction (**Supplemental Figure 2, Supplemental Table 2**). Thus, elevated BMI is associated with specific cancer genotypes independent of genetic ancestry, sex, and age.

We further considered the possibility that other demographic factors, etiologies, or exposures might confound an association between BMI and tumor genotypes. To directly examine this possibility we focused on lung adenocarcinoma, where there is an established association between smoking status and elevated frequency of *KRAS* mutations/reduced frequency of *EGFR* mutations^11^. We modeled the presence of driver mutations in *KRAS* and *EGFR* as a function of BMI, smoking status, ancestry, age, and sex, for 4,150 lung adenocarcinoma patients with adequate genomic and clinical data for this analysis. The association between BMI and EGFR (p *=* 4.5×10^−4^ estimate = −0.03) and BMI and *KRAS* (p-value = 1.6×10^−3^ estimate = 0.02) remained statistically significant after controlling for all covariates (**Figure 1b and 1c, Supplemental Table 3**). Thus, high BMI is associated with elevated rates of *KRAS* mutations and lower rates of *EGFR* mutations in lung adenocarcinoma independent of smoking status.

To corroborate these findings in an independent cohort, we sought to replicate the two associations with the strongest evidence in our dataset: *EGFR* and *KRAS* mutations in lung adenocarcinoma. We obtained somatic mutation calls and BMI information from 2,727 lung adenocarcinoma patients at a separate institution. Analysis of this dataset confirmed that *EGFR* mutations were negatively associated with BMI (p = 5×10^−2^ estimate = −0.02) and *KRAS* mutations were positively associated with BMI (p = 9×10^−4^ estimate = 0.03) (**Fig 1e, Supplemental Table 4**), after controlling for age, sex, and ancestry. Thus, in lung adenocarcinoma, obesity appears to independently predispose patients to higher frequency of *KRAS* mutations, and lower frequency of *EGFR* mutations.

Finally, we considered the possibility that associations between even early BMI measurements and genomic alterations were indirectly induced by cancer-associated weight changes such as cachexia. To evaluate this possibility in lung cancer, we reviewed initial diagnosis medical notes from a subset of treatment naive lung adenocarcinoma patients treated at MSK and classified them according to whether they exhibited signs of cachexia, anorexia, or other unexplained weight loss in the six months prior to diagnosis (see **Methods**). There was no association between clinically notable weight loss prior to diagnosis and the presence of *EGFR* (p-value = 0.70 odds ratio = 1.16) or *KRAS* (p-value = 0.69 odds ratio = 1.15) mutations (**Figure 1f**), indicating that the association between BMI and these mutations is not confounded by pre-diagnosis weight loss.

Previous work has demonstrated the modulatory effects of obesity on the emergence of cancer, which is complex and results in higher presence of certain cancers such as liver and colorectal and lower frequency of others such as lung cancer. Our results suggest that, after controlling for numerous host and environmental factors, obesity predisposes patients even more specifically to cancers with particular tumor genotypes.

Our data suggests that *EGFR*-mutant lung adenocarcinoma has a disproportionately lower mutation frequency in obese patients, and conversely that *KRAS-*mutant lung adenocarcinoma exhibits an elevated mutation frequency in obese patients. These findings currently have treatment implications, as a broader portion of *EGFR* mutations are amenable to targeted therapy than *KRAS*, although this landscape is changing rapidly^12^. More fundamentally, few modifiable risk factors of *EGFR/KRAS*-mutant lung adenocarcinoma have been characterized. A recent study^6^ suggests that air pollution may trigger microenvironmental changes allowing for proliferation of cells with this activated oncogenic driver. This is the first study suggesting that obesity is a similar modifier of *EGFR*-mutant lung adenocarcinoma risk. There are many plausible mechanisms by which obesity might mediate this effect, including *EGFR-* and *KRAS-*mediated modulation of lipid metabolism in cancer cells via PIK3CA/mTOR pathways^7^. Obesity itself may also inhibit the function of CD8+ T cells^5^, which can affect the selective pressure for immunogenic driver alterations. Because *KRAS*-mutant lung adenocarcinoma is generally more sensitive to immune checkpoint blockade than *EGFR*-mutant lung adenocarcinoma^13^, it seems plausible that changes to the immune microenvironment downstream of obesity may disproportionately allow for proliferation of *KRAS-*mutant lung adenocarcinoma in patients with obesity.

Our study has several limitations. We were underpowered to detect associations between BMI and genomic alterations in rare cancer types and infrequently mutated genes. There may be other genomic changes, including focal copy number alterations and structural rearrangements, associated with BMI which we did not consider in this analysis. BMI itself is only one of many metrics to study obesity, and we did not have access to such other metrics in the clinical record^3^. Genomic studies do not fully capture the molecular landscape of a tumor, and broader studies that combine genomic, transcriptomic, and metabolomic analyses would provide a more well-rounded view of the effect of obesity on cancer tumor phenotypes^14^. Finally, and most importantly, prospective studies will have to be done to determine whether modifications to BMI, either through diet, exercise, or medical interventions, can causally modify the incidence of particular cancer genotypes.

## Methods

### Patient Cohort

MSK-IMPACT (NCT01775072), a prospective observational cohort study of tumor evolutions, was approved by the Institutional Review Board at Memorial Sloan Kettering Cancer Center. Patients provided written informed consent for the use of their genomic data for research. Participants were not compensated for their participation. Tumors were sequenced using the MSK-IMPACT assay through 23/03/2023. In patients with multiple samples, only one sample (the earliest sampled primary tumor) was included in the final cohort. Clinical characteristics were annotated per the standard MSK-IMPACT workflow. The total cohort constitutes 34,274 samples spanning 102 cancer types. Only cancer types with over 50 samples were tested for genotype-BMI associations. For each patient, we identified the BMI measurement collected at the date nearest to the date of sample acquisition leading to genomic sequencing, i.e. the earliest possible date for which a given genomic alteration could be confirmed in a patient’s tumor. Patients without a BMI measurement within 30 days of tumor acquisition were excluded. Patients at Dana Farber Cancer Institute had tumor genomic profiling with Oncopanel^15^, a targeted sequencing platform analogous to MSK-IMPACT.

### Targeted DNA Sequencing with MSK-IMPACT

DNA sequencing was performed using the MSK-IMPACT sequencing panel, which is a hybridization capture-based next-generation sequencing assay, in a Clinical Laboratory Improvement Amendments (CLIA)-certified molecular laboratory. Genomic DNA from formalin-fixed paraffin-embedded (FFPE) primary or metastatic tumors and matched normal samples was extracted and sheared. Custom probes were then synthesized for targeted sequencing of all exons and select introns of 341, 410, 468, or 505 genes. Illumina HiSeq 2500 was used to capture pooled libraries containing captured DNA fragments to high, uniform coverage (x>500 median coverage). All classes of genomic alterations including substitutions, indels, copy number alterations, and rearrangements were determined and called against the patient’s matched normal sample. The computational pipelines used for variant calling are based on standard best practices and use a combination of open-source and custom-written scripts and programs.

The OncoKB precision oncology knowledge base was used to annotate genomic alterations. OncoKB identifies functionally relevant cancer variants and their potential clinical actionability. Only alterations classified as oncogenic by OncoKB were used in this analysis. Reported alteration frequencies were adjusted to account for the specific set of genes included in each version of the MSK-IMPACT panel used by dividing the number of gene-specific alterations by the number of samples for which a given gene was sequenced. Somatic alterations were labeled as therapeutically actionable using levels of clinical actionability defined in OncoKB. These levels range from Level 1, FDA-approved biomarkers of response to FDA-approved drugs, to Level 4, biomarkers of hypothetical relevance based on compelling preclinical biological evidence.

### Logistic Regression Model

Logistic regression was performed where BMI (as a continuous variable) was used to predict somatic mutation status (“Gene”) in a particular cancer type. Only detailed cancer types with over 50 patients and the 341 genes in the initial MSK-IMPACT panel were included in this study. Covariates for the logistic regression included age, sex, and ancestry. Smoking status was included in models with lung adenocarcinoma. The glm function from the stats R package was used to conduct the modeling. P-values were Benjamini-Hochberg corrected.

The following equations were used.

1. Gene ∼ BMI
2. Gene ∼ BMI + age + sex + ancestry
3. Gene ∼ BMI + age + sex + ancestry + smoking status

### Pre-diagnosis weight loss identification and analysis

To identify pre-diagnosis weight loss in patients, 800 initial consultation notes were randomly selected from patients with treatment naive lung adenocarcinoma and were manually categorized. Records with mentions of the patient reporting weight loss, anorexia, or decreased appetite were labeled as having pre-diagnosis weight loss.

## Data Availability

All clinical and genomic sequencing data described in this manuscript has been deposited in the cBioPortal for Cancer Genomics and are available for online browsing and download there. While raw sequencing data is restricted to protect patient privacy in accordance with federal and state law, de-identified data are available. De-identified data can be requested for research use from the corresponding authors. Data will be shared for a span of 2 years within 2 weeks of execution of a data transfer agreement with MSKCC, which retains all title and rights to the data and results from their use.

## Code Availability

Code for reproducing the results in Figure 1 and associated Supplementary Tables is available at https://github.com/reznik-lab/bmi_genomics.

## Acknowledgements

We thank the members of the Reznik, Jee, and Schultz laboratories for discussion and support. This work was supported by P50 CA247749-01, P30 CA008748, the Molecular Diagnostics Service in the Department of Pathology, and the Marie-Josee and Henry R. Kravis Center for Molecular Oncology. J.J. was supported by K12 CA184746. E.R. was supported by NIH R37 CA276200 and DOD grant HT9425-23-1-0995.

## Author contributions

E.R. and J.J. conceived the study. C.T., M.W.C., C.F., T.P., V.J.C., S.K., N.S., I.O., and A.G. assisted with genomic data collection and analytical methodology development. C.T., V.J.C., A.G., J.J., and E.R. designed and performed the experiments. C.T., V.J.C., J.J., and E.R. wrote the manuscript with input from all authors.

## Competing financial interests

E.R. is a paid consultant of Xontogeny, LLC.

## Supplementary Materials

**Fig. S1:**
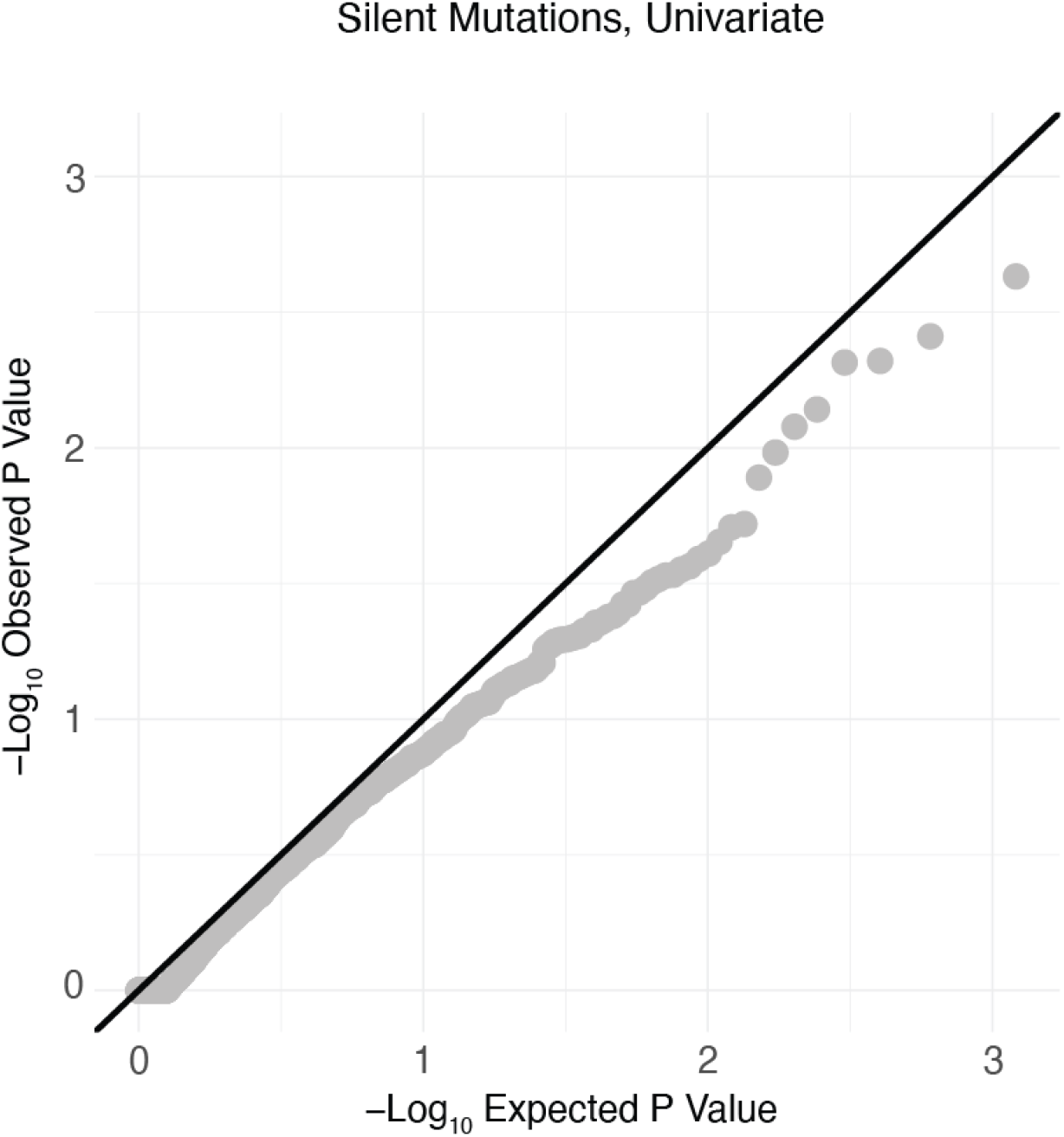
Silent mutations are not associated with body mass index. Scatterplot comparing −log_10_ observed p-values vs. −log_10_ observed p-values in univariate logistic regression models for silent mutations.

**Fig. S2:**
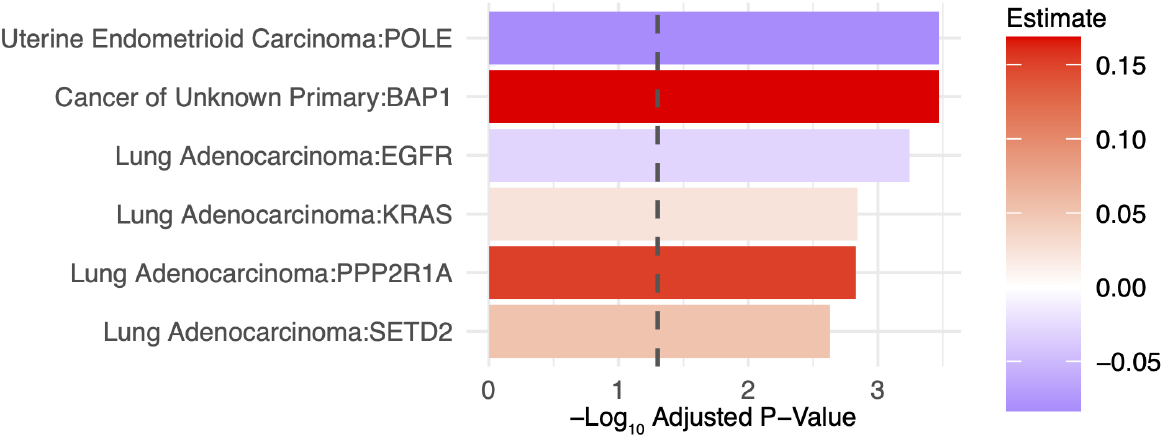
Association between BMI and mutation incidence is preserved after controlling for clinical covariates. Barplot of −log_10_ p-values for significant gene:cancer type pairs from multivariate logistic regression models.

## Notes

### Competing Interest Statement

ER is a paid consultant of Xontogency LLC.

### Author Declarations

All patients provided written informed consent and were prospectively sequenced as part of their active care at Memorial Sloan Kettering Cancer Center (MSKCC) as part of an Institutional-Review-Board-approved research protocol (NCT01775072).

### Summary of Updates

Corrected name of co-author Alexander Gusev.

